# Real-time RT-PCR Allelic Discrimination Assay for Detection of N501Y Mutation in the Spike Protein of SARS-CoV-2 Associated with Variants of Concern

**DOI:** 10.1101/2021.06.23.21258782

**Authors:** Mariana Abdulnoor, AliReza Eshaghi, Stephen J. Perusini, Antoine Corbeil, Kirby Cronin, Nahuel Fittipaldi, Jessica D. Forbes, Jennifer L. Guthrie, Julianne V. Kus, Anna Majury, Tony Mazzulli, Roberto G. Melano, Romy Olsha, Ashleigh Sullivan, Vanessa Tran, Samir N. Patel, Vanessa G. Allen, Jonathan B. Gubbay

## Abstract

The N501Y amino acid mutation caused by a single point substitution A23063T in the spike gene of SARS-CoV2 is possessed by the three most common variants of concern - B.1.1.7, B.1.351, and P.1. A rapid screening tool using this mutation is important for surveillance during the COVID-19 pandemic.

We developed and validated a single nucleotide polymorphism real-time reverse transcription polymerase chain reaction assay using allelic discrimination of the spike gene N501Ymutation to screen for potential variants of concern and differentiate them from wild-type SARS-CoV-2. A total of 160 clinical specimens positive for SARS-CoV-2 were characterized as mutant (N501Y) or wild-type by Sanger sequencing and were subsequently tested with the N501Y single nucleotide polymorphism real time reverse transcriptase polymerase chain reaction assay. Our assay compared to sequencing, the gold standard for SNP detection and lineage identification, demonstrated clinical sensitivity of 100% for all 57 specimens displaying N501Y mutant, which were confirmed by Sanger sequencing to be typed as A23063T, including one specimen with mixed signal for wildtype and mutant. Clinical specificity was 100% in all 103 specimens typed as wild-type, with A23063 identified as wild-type by Sanger sequencing. The identification of circulating SARS-CoV-2 lineages carrying an N501Y mutation is critical for surveillance purposes. Current identification methods rely primarily on Sanger sequencing or whole genome sequencing which are time-consuming, labor-intensive and costly. The assay described herein is an efficient tool for high-volume specimen screening for SARS-CoV-2 VOCs and for selecting specimens for confirmatory Sanger or whole genome sequencing.

## Introduction

Severe acute respiratory syndrome coronavirus 2 (SARS-CoV-2) is the viral etiology of the coronavirus disease 2019 (COVID-19) pandemic. Several SARS-CoV-2 variants of concern (VOC) have been identified which are associated with increased transmissibility, increased virulence, changes in clinical disease presentation and decreased effectiveness of public health measures, diagnostics, vaccines, and therapeutics (1). In 2020, three novel VOCs of SARS-CoV-2 emerged independently (2-4).

The VOC first detected in the UK represents the B.1.1.7 lineage (also known as 20I/501Y.V1 and VOC202012/010) and contains three key mutations in S gene 69-70del, N501Y and P681H. Recently, E484K has been observed in some B.1.1.7 genome sequences (5). B.1.1.7 is associated with increased transmissibility of 50-70% (6), and increased disease severity and risk of death (7). Cases of B.1.1.7 lineage have been found in 114 countries (8). In Ontario, Canada, 2,165 cases of B.1.1.7 have been confirmed as of April 5, 2021(9).

A second SARS-CoV-2 VOC representing the B.1.351 lineage (also known as 20H/501Y.V2 and VOC-202012/02) emerged in October 2020 in South Africa, and rapidly has become the dominant circulating strain in that country (3). This VOC has multiple mutations in the receptor binding domain (RBD) of the S protein, including K417N, E484K, and N501Y. The B.1.351 lineage is associated with increased transmissibility (10) and immune evasion (11,12). It is unknown how this VOC influences disease severity, hospitalizations and deaths. B.1.351 has been detected in 68 countries (13) including Canada, where in Ontario there are 71 confirmed cases, as of April 5, 2021(9).

A third emerging VOC is the P.1 lineage (also known as B.1.1.28.1, 20J/501Y.V3, and VOC-202101/02), first detected in Japan from travelers returning from Brazil (14). This VOC shares the S gene N501Y single nucleotide polymorphism (SNP) with B.1.1.7 and the E484K SNP with B.1.351; however, all three lineages arose independently (4). The P.1 lineage may have higher inherent transmissibility compared to previous lineages (14). This lineage is associated with antigenic escape (12). The P.1 lineage has been identified in 36 countries (15) including Canada, where there are 106 confirmed cases in Ontario as of April 5, 2021 (9).

These SARS-CoV-2 VOCs will likely hamper public health efforts to contain the spread of the virus. Thus, surveillance is important to control the spread of these lineages. Current methods rely on genome sequencing, either by Sanger sequencing or whole genome sequencing (WGS). These approaches, although of high accuracy, are labour-intensive and are not amenable to rapid turnaround times as they require extensive laboratory processing and bioinformatic analysis. They usually take several days to complete, therefore testing capacity is quickly outpaced as the number of specimens to be analyzed increases with transmission.

The use of real-time reverse transcription PCR (rRT-PCR) to detect SNPs of relevance would facilitate rapid preliminary identification of potential VOCs and initiation of public health measures. Herein, we report a laboratory developed SNP rRT-PCR protocol to detect the presence of N501Y, which can act as a screening tool for the three circulating VOCs in Ontario as they all share this SNP. Specimens with N501Y detected are subsequently submitted for WGS to confirm the presence of a VOC and identify the SARS-CoV-2 VOC lineage.

## Methods

### Clinical Specimens

Public Health Ontario (PHO) Laboratory is the reference microbiology laboratory for the province of Ontario, Canada. As part of the response to detect VOCs in Ontario, PHO Laboratory developed indications for VOC screening, which were similar to Canadian guidelines (16). Clinical specimens that met these criteria with a cycle threshold (Ct) value ≤30 underwent Sanger sequencing of a 698 bp (nucleotide positions 22516 to 23214) S gene fragment, which includes the locations of key RBD mutations (K417N/T, E484K, N501Y). Specimens with mutations detected underwent further sequencing to confirm if a VOC was present. In addition, a subset of all SARS-CoV-2 positive specimens underwent WGS as part of the provincial genomic surveillance program, which included VOC screening.

A convenience sample of 160 SARS-CoV-2 positive clinical specimens collected between November 20, 2020 and January 20, 2021, submitted to PHO Laboratory from across the province, that had undergone VOC screening by partial S gene sequencing, as described above, were used in the optimization and validation of the assay. No clinical information was available when sequencing was conducted. However, the presence of N501Y was known prior to running specimens in our assay as it was previously sequenced. All SARS-CoV-2 positive specimens with N501Y available at PHO Laboratory were included. Specimen types included nasopharyngeal swabs, throat swabs, and bronchoalveolar lavage fluid.

### rRT-PCR Design and Optimization

Primer and probe sequences were selected using an alignment of spike nucleotide sequences of a wild-type SARS-CoV2 reference genome (GenBank accession: MN908947.3) and a representative of the VOC B.1.1.7 lineage (GSAID accession: EPI_ISL_601443). Selected primer and probe sequences were aligned to 9000 gene sequences downloaded from GISAID with submission date between December 1, 2020 through December 6, 2020 to ensure they were targeting a conserved region. In addition, a BLAST search of the primer and probe sequences against the whole genome of the reference sequence (MN908947.3) was conducted to ensure the absence of homology to other parts of the SARS-CoV-2 genome. Primers and probes selected for the detection of SNP A23063T are reported in **Supplementary Table 1**. Primer and probes were synthesized by LGC, Biosearch Technologies (Middlesex, United Kingdom). Optimization was performed using previously characterized wild-type and B.1.1.7 specimens. No B.1.351 or P.1 lineage SARS-CoV-2 were detected in Ontario at the time of assay development.

An additional smaller validation was conducted using primers and probes from a second vendor, Thermo Fisher Scientific (Massachusetts, United States), to allow redundancy of reagent suppliers.

### Total Nucleic Acid Extraction

Specimens underwent nucleic acid extraction using either Perkin Elmer chemagic™ 360 automated systems (PerkinElmer, Massachusetts, United States) (input volume 300 µl; eluate volume 60 µl) or MGISP-960RS platform (MGI Tech Co., Guangdong, China) (input volume 180 µl; eluate volume 45 µl) according to the manufacturer’s recommendations.

### N501Y SNP rRT-PCR Assay

The PCR amplification was performed using the Taqpath™ 1-Step Multiplex Master Mix on the Applied Biosystems™ QuantStudio™ 5 Real-Time PCR system (Thermo Fisher Scientific, Massachusetts, United States).

A total volume of 10µl reaction mix contained 2.5µl of 4X TaqPath™ 1-Step Multiplex Master Mix, 1.0µl of primer mix, 1.0µl of probe mix, 2.5µl of RNase-free water, and 3µl of viral RNA extract. The default thermocycling profile suggested for 4X TaqPath™ 1-Step Multiplex Master Mix was used including 20 minutes at 25° C, 10 minutes at 53°C, 2 minutes at 95°C and 45 cycles of 95°C for 3 seconds then 60°C for 30 seconds. The controls used in each run were an extraction negative control that consisted of a nuclease-free water processed along with SARS-CoV-2 specimens during the extraction process, a PCR negative control that consisted of nuclease-free water in place of nucleic acid in the reaction mix, and a positive RNA amplification control consisting of combined wild-type and B.1.1.7 SARS-CoV-2 RNA obtained from clinical specimens positive by in-house end-point PCR and sequencing. During optimization, it was determined that nonspecific signals occurred above a Ct of 37, therefore only results with Ct ≤37 were considered positive for either probe target. The assay does not have an indeterminate range. The time interval between sequencing and running the N501Y rRT-PCR assay ranged from one to 14 days.

### PCR Amplification and Sequencing of Partial Spike Gene

Sanger sequencing was chosen as the comparator method as it is a highly accurate methodology to determine the presence of SNPs, is the gold standard for identifying SNPs, and was the method in use at PHO Laboratory for VOC detection. A detailed explanation of the Sanger sequencing protocol used is provided in **Supplementary appendix A**. Specimens with Ct values ≤35 with a good quality Sanger chromatogram, containing evenly-spaced peaks each with one colour, and the least amount of baseline noise, were considered positive. Sanger sequencing failed when Ct values were >35 and the chromatogram contained mixed bases and was messy.

### Validation

Primary outcome measures were clinical sensitivity, clinical specificity, and precision.

### Accuracy – Clinical Sensitivity and Specificity

A panel of 160 SARS-CoV-2 positive clinical specimens that had been characterized by partial Sanger sequencing of the S gene were used for validation of the N501Y SNP assay. The result obtained by our rRT-PCR N501Y SNP assay was compared to the known sequence results previously obtained by partial S gene sequencing and used to calculate sensitivity and specificity. Sensitivity of >90% and specificity >95% were considered acceptable.

### Intra-run repeatability

Intra-run repeatability was evaluated by running three replicates of 10 N501Y VOCs (including 1 with a mixed population) and 5 wild-type samples in the same rRT-PCR run and comparing Ct values across the replicates. The threshold for acceptance between runs was a Ct difference of <3.3.

### Inter-run reproducibility

Inter-run reproducibility was evaluated by rerunning the panel used for intra-run repeatability on a second day. This panel was also sent out to six other microbiology laboratories in Ontario to validate the assay prior to implementation. The threshold for acceptance was >95% agreement between labs.

### Ethical Statement

The PHO Ethics Review Board has determined that this project did not require research ethics committee approval, as it describes analyses that were completed at PHO Laboratory as part of routine clinical respiratory testing and surveillance during the COVID-19 pandemic in Ontario and are, therefore, considered public health practice and exempt from this requirement.

## Results

### Accuracy - Clinical Sensitivity and Specificity

To analyze the performance of the N501Y SNP assay, a panel of 160 specimens including 57 specimens with N501Y detected by Sanger sequencing, and 103 wild-type were tested. One of the 57 N501Y positive specimens contained a mixed result on Sanger sequencing, indicating the presence of both wild-type and mutant strains. All specimens with a known A23063T substitution in the Sanger chromatogram produced a positive signal for N501Y with the rRT-PCR SNP assay (57/57, 100%), and were confirmed to be B.1.1.7 lineage by Sanger sequencing analysis. All samples with known A23063 wild-type characterization by Sanger showed positive signal for wild-type with the rRT-PCR SNP assay (103/103, 100%). The positive predictive value was 100%% (57/57) and the negative predictive value was 100% (103/103) (**Table 1)**.

**Table 1:** Summary of accuracy testing data for PHO Laboratory SARS-CoV-2 N501Y SNP rRT-PCR assay

|  | **Sanger**<br>**Mutant** | **Sanger Wild-type** | **Total** |
| --- | --- | --- | --- |
| **SNP (N501Y) detected** | 57 | 0 | 57 |
| **SNP (N501) not**<br>**detected** | 0 | 103 | 103 |
| **Total** | 57 | 103 | 160 |

There was one specimen with a mixed population of wild-type and mutant, this specimen was counted as positive for N501Y.

**Table 2:**
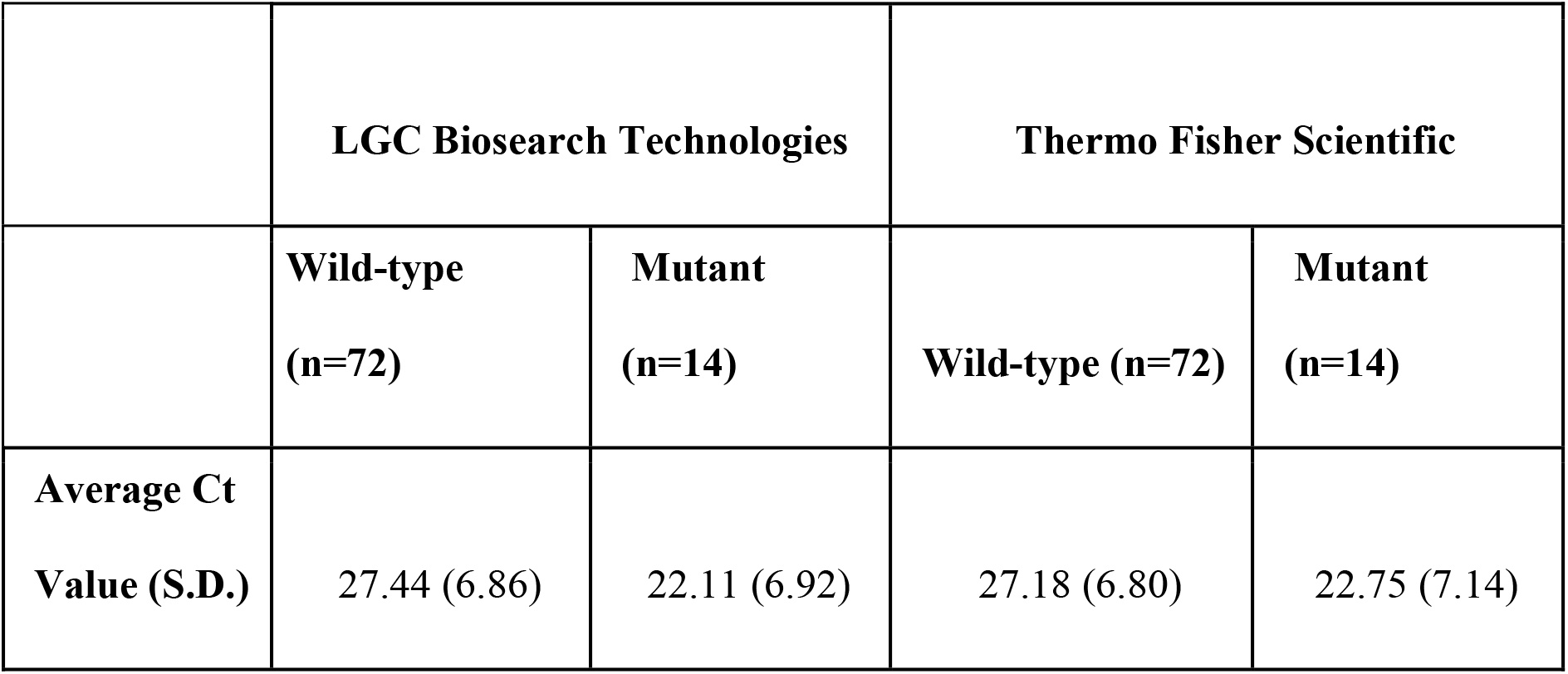
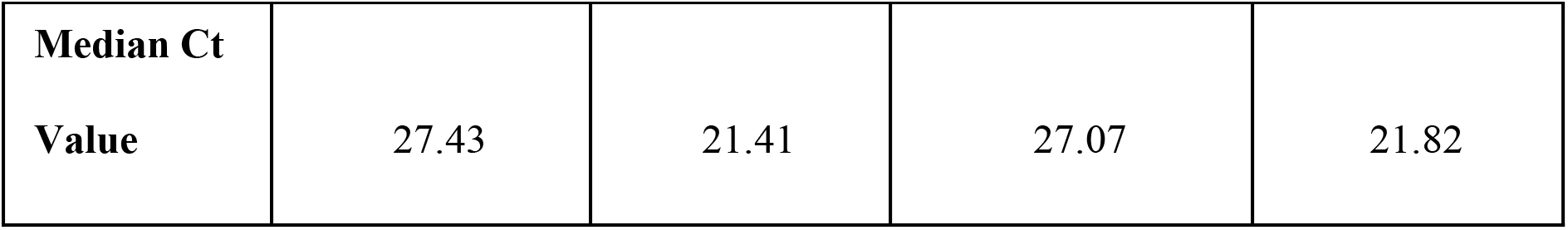
Results of validation experiments with primer and probe sets from two vendors, LGC Biosearch Technologies and Thermo Fisher Scientific.

The mean E gene and N501Y SNP rRT-PCR assay Ct values of all VOC specimens in this study were 19.15 (S.D. 3.76) and 21.88 (S.D. 3.80), respectively. The mean Ct values of E gene and N501 SNP rRT-PCR for wild-type specimens were 20.87 (S.D. 5.38) and 23.75 (S.D. 5.52), respectively. (**Supplementary Table 2**).

### Comparison of two primer and probe sets

Our N501Y SNP assay was validated using primer and probe sets from two vendors, LGC, Biosearch Technologies (Middlesex, United Kingdom) and Thermo Fisher Scientific (Massachusetts, United States). A subset of 87 specimens, including 14 mutants, 72 wild-type and one mixed population were tested (**Table 1**). We observed comparable results for both primer and probe sets, as there were no significant differences in average Ct values or number of N501Y specimens detected.

### Precision – intra-run repeatability and inter-run reproducibility

The analysis of intra-run repeatability was done with a panel of nine N501Y samples, five wild-type, one mixed N501Y/N501 sample, and five SARS-CoV-2 negative samples. The panel was run on three replicates. The results show 100% agreement (**Supplementary Table 3**) with an average Ct 26.17 (average SD 0.19, average coefficient of variation 0.71%) for the mutant specimens and average Ct 26.76 (average SD 0.16, average coefficient of variation 0.6%) for wild-type specimens.

The analysis of inter-run reproducibility was done with replicates of a panel of nine N501Y samples, five wild-type samples, one mixed N501Y/N501 sample, and five negative samples. Among the 10 runs obtained from seven laboratories, the results showed 100% agreement with an average Ct 26.04 (SD 0.98, coefficient of variation 4%) for mutant specimens and an average Ct 27.39 (SD 1.29, coefficient of variation 5%) for wild-type specimens.

## Discussion

Here we describe the validation of a N501Y SNP rRT-PCR assay to provide rapid screening of SARS-CoV-2 positive specimens for VOCs. Currently, three VOCs (B1.1.7, B.1.351 and P.1) are circulating in Ontario and surveillance of these VOCs is of considerable public health importance. The World Health Organization recommends VOC surveillance to help control the spread of VOCs (1). In our workflow, positive SARS-COV-2 specimens are screened for the N501Y SNP using the described rRT-PCR assay then potentially submitted for WGS to confirm VOC identification and determine the VOC lineage. There were no adverse events from performing the N501Y SNP rRT-PCR assay as individuals had already been classified as having the mutation or not based on previous sequencing. Our assay is efficient, and clinically validated with a high degree of sensitivity and specificity for detection of the N501Y SNP.

Sanger sequencing or WGS of VOCs require longer turnaround times, are expensive, and some regions may not have access to sequencing facilities or the ability to scale-up their sequencing capabilities. A two-factor screening process for B.1.1.7 detection has been employed to aid with surveillance efforts (17). The protocol detects the 69-70del in the S gene of B.1.1.7 as this deletion causes S-gene target failure (SGTF) in certain assays (e.g. Thermo Fisher’s TaqPath COVID-19 Combo Kit). Once SGTF is detected, specimens are sequenced by WGS. SGTF has been used as a proxy for the B.1.1.7 VOC (6). This method is limited to detecting VOCs carrying 69-70del. A multiplex rRT-PCR to screen for VOC was published by Vogel et al. (18) that targets the aa3675-3677 deletion in ORF1a gene as well as the aa69-70del in the S gene. The ORF1a deletion of aa3675-3677 is present in B.1.17, B.1.351, and P.1 and the aa69-70del in S gene is used to differentiate B.1.17 from B.1351 and P.1. However, the ORF1a deletion is not found in all B.1.351 VOCs, as there is a monophyletic clade that does not contain this deletion. Another group (19) published a rRT-PCR assay to detect only the N501Y SNP and requires a melting curve to confirm the specimens as wild-type if the test is negative. Our procedure differs as it is able to detect both the wild-type and the SNP, avoiding the additional step of analyzing melting-curves and thus providing easier interpretation of the results.

To understand the provincial prevalence of VOCs in Ontario, the majority of SARS-CoV-2 positive specimens reported on January 20, 2021 across the province were sent for screening using our rRT-PCR N501Y SNP assay (20). Our rRT-PCR N501Y SNP assay has now been instituted for surveillance testing across Ontario, allowing notification to public health at least one to seven days earlier than relying on Sanger sequencing or WGS. On March 11, 2021, 1,138 specimens were screened with our N501Y rRT-PCR assay, 897/899 (99.8%) specimens that tested positive for N501Y SNP were confirmed through WGS and 238/239 (99.6%) were confirmed wildtype through WGS (data extracted from PHO laboratory information system). Using this assay, we’ve now documented a rapid rise in VOC cases with roughly 60% of cases on April 5, 2021 identified as VOC or containing N501Y and/or E484K SNPs (9).

Because our assay only detects the N501Y SNP, new emerging lineages that do not carry this SNP, such as B.1.525, will be missed (21). The assay is also unable to differentiate between VOCs that share the N501Y SNP; therefore, an additional sequencing step is required to delineate the lineage. An additional SNP assay target detecting E484K was incorporated into this assay in a multiplex format as of March 22, 2021, and assays to detect E417T/N S gene mutations are currently under development in our laboratory as these could provide further characterization of currently known VOCs without requirement of WGS. Even with additional targets, definitive lineage identification is not possible based on SNP assays alone and requires sequencing. Our validation was conducted with specimens characterized as B.1.1.7 but when implemented as a screening tool we were able to detect B.1.351 and P.1 VOCs. Our validation dataset contained a small sample size of 57 positive specimens; however, clinical sensitivity and specificity were further confirmed in our data review post-implementation. A limit of detection was not performed, however, comparison of rRT-PCR SNP Ct values to E gene Ct values using the Corman et al (22) assay, shows the SNP assay is approximately 1 log less sensitive (3-4 cycles higher). We did not conduct analytical specificity tests with a cross reactivity panel due to limited availability of SARS-CoV-2 VOC strains. In our analysis we did not assess baseline demographics, clinical characteristics of the patients and severity of disease, as limited information is provided to PHO Laboratory, which provides testing as a reference laboratory. Our assay is not intended for diagnostic purposes, but rather as a screening tool to aid with faster identification of VOCs through differentiation of N501 and N501Y.

Our assay is a quick and simple tool that can be applied to screening SARS-CoV-2 positive specimens from priority groups at greater risk for VOC infection such as international travellers, suspected SARS-CoV-2 reinfection, infection post COVID-19 vaccination, and outbreaks. Ontario is screening all positive specimens, which can be in excess of 1000 specimens per day, and the N501Y SNP rRT-PCR assay has allowed us to screen a high volume of specimens. Due to high specificity and positive predictive value, any specimen that is N501Y positive can be presumed to be a VOC while awaiting characterization by WGS. Early, high-throughput screening with PHO Laboratory’s N501Y screening assay allows effective public health measures to be put in place to limit the spread of VOCs in the community.

## Data Availability

Data requests can be made to the corresponding author for institutional review.

## Acknowledgement

We acknowledge the collaboration of members of the Ontario Provincial COVID-19 Diagnostic Network. We acknowledge the staff of Virus Detection and Molecular Diagnostics, Public Health Ontario Laboratory, for diagnostic testing and N501Y screening of SARS-CoV-2 specimens. We thank the Public Health Ontario Laboratory research group for conducting SARS-CoV-2 whole genome sequencing.

---

**Supplementary Table 1:**
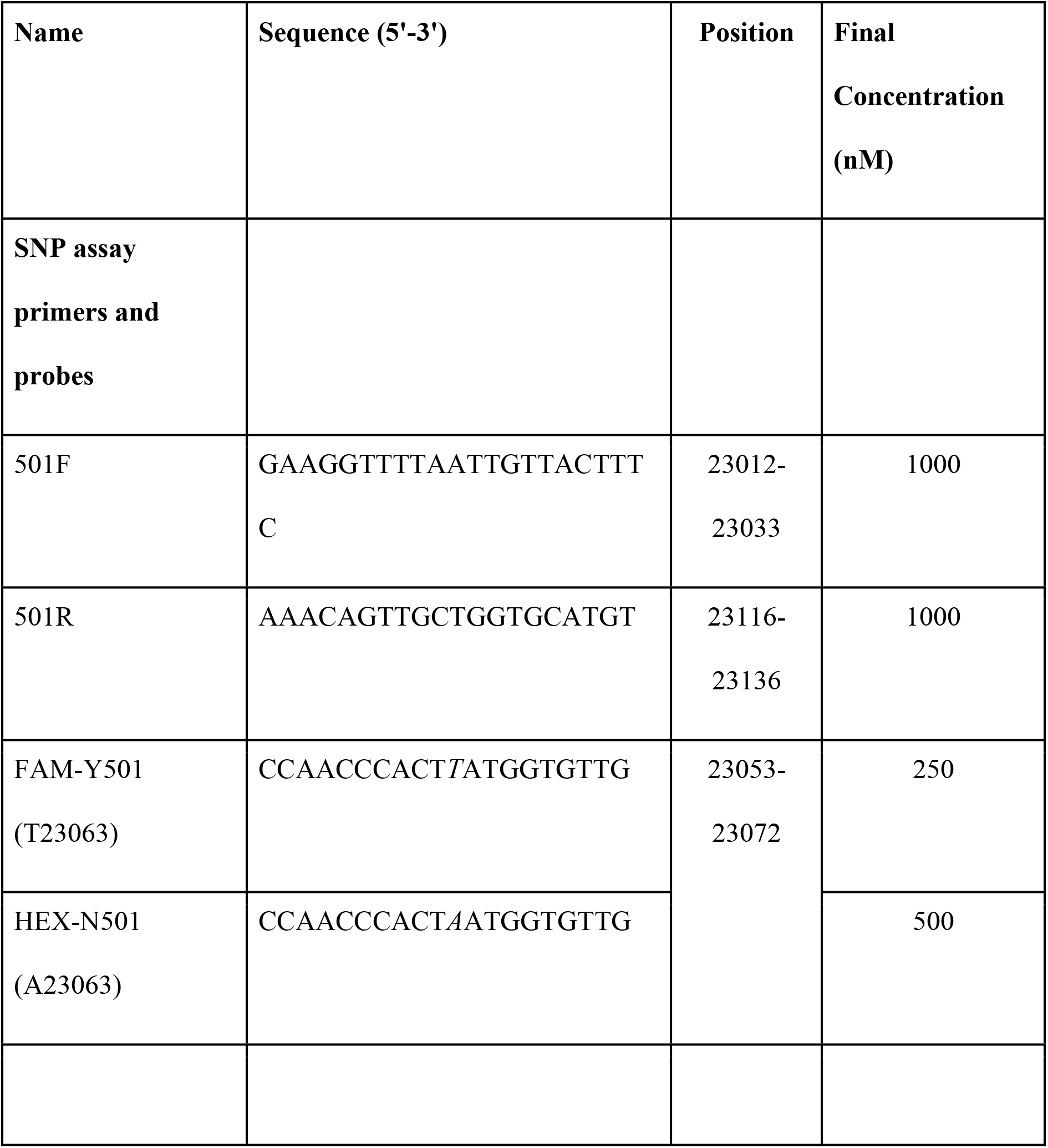
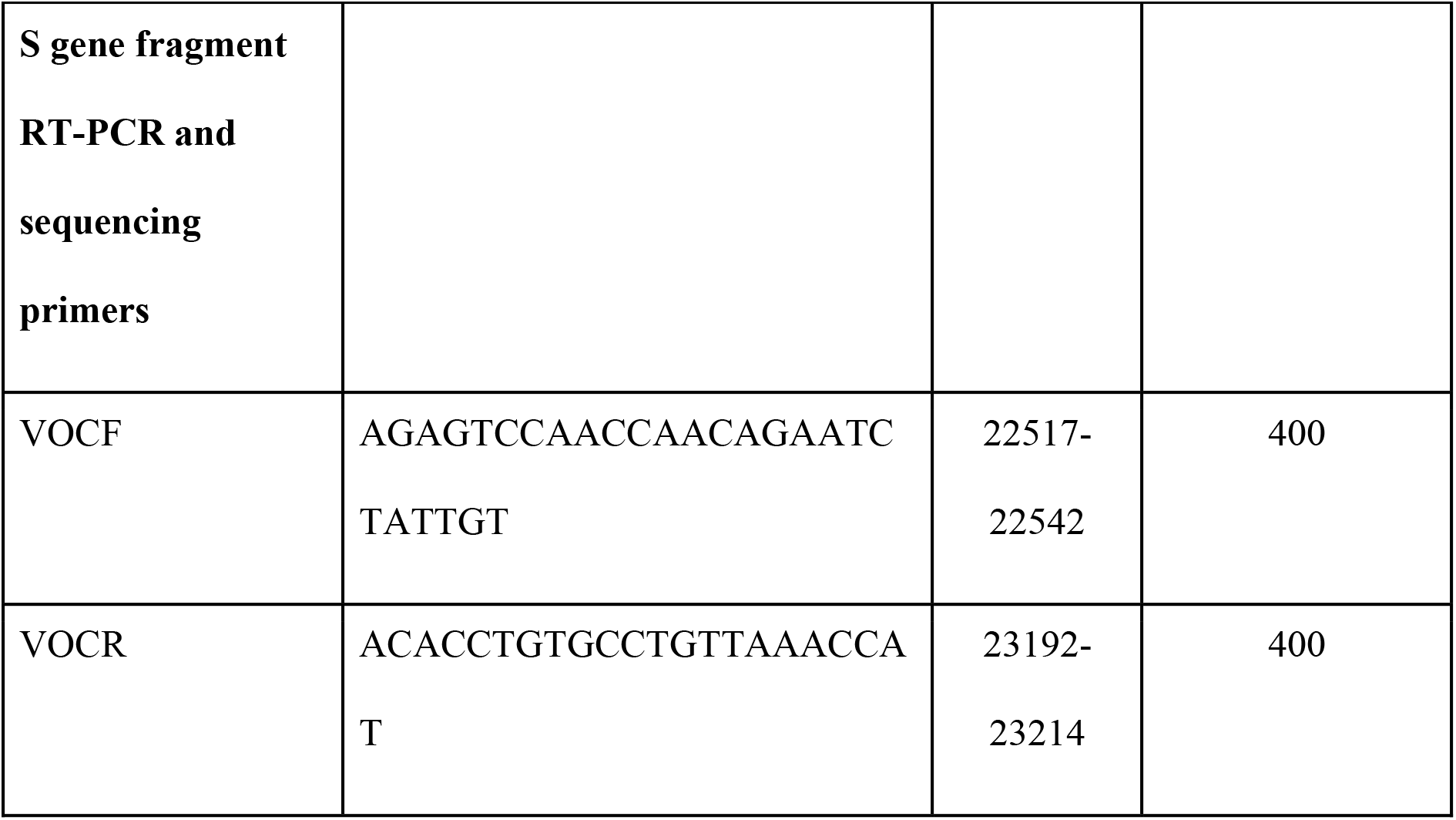
Primers and probes used in the PHO Laboratory SARS-CoV-2 N501Y SNP rRT-PCR assay and Sanger sequencing

**Supplementary Table 2:** Individual specimen cycle threshold (Ct) values for E gene (22) and PHO Laboratory rRT-PCR N501Y assay.

| Specimen ID | E gene<br>Ct | SNP assay |  | SNP<br>results | Sanger Sequencing |  |  | Result |
| --- | --- | --- | --- | --- | --- | --- | --- | --- |
|  |  |  |  |  | K417N | E484K | N501<br>Y |  |
|  |  | N501 | N501<br>Y |  | G2281<br>3T | G2301<br>2A | A2306<br>3T |  |
| 1 | 18.81 | 22 | ND<sup>a</sup> | Neg | G | G | A | Wild-type |
| 2 | 22.93 | 27 | ND | Neg | G | G | A | Wild-type |
| 3 | 23.33 | 26.9 | ND | Neg | G | G | A | Wild-type |
| 4 | 15.08 | 17.8 | ND | Neg | G | G | A | Wild-type |
| 5 | 18.16 | 21.6 | ND | Neg | G | G | A | Wild-type |
| 6 | 19.37 | 22.3 | ND | Neg | G | G | A | Wild-type |
| 7 | 13.04 | 16.7 | ND | Neg | G | G | A | Wild-type |
| 8 | 15.03 | 19 | ND | Neg | G | G | A | Wild-type |
| 9 | 24.14 | 27.7 | ND | Neg | G | G | A | Wild-type |
| 10 | 29.78 | 33.4 | ND | Neg | G | G | A | Wild-type |
| 11 | 19.22 | 22.9 | ND | Neg | G | G | A | Wild-type |
| 12 | 28.95 | 32.1 | ND | Neg | G | G | A | Wild-type |
| 13 | 20.24 | ND | **24.9** | **Pos** | G | G | **T** | N501Y |
| 14 | 15.73 | ND | **20.4** | **Pos** | G | G | **T** | N501Y |
| 15 | 18.29 | ND | **22.2** | **Pos** | G | G | **T** | N501Y |
| 16 | 19.00 | ND | **22.9** | **Pos** | G | G | **T** | N501Y |
| 17 | 18.24 | ND | **22.5** | **Pos** | G | G | **T** | N501Y |
| 18 | 22.59 | ND | **26.7** | **Pos** | G | G | **T** | N501Y |
| 19 | 18.97 | ND | **22.7** | **Pos** | G | G | **T** | N501Y |
| 20 | 14.6 | 18.3 | ND | Neg | G | G | A | Wild-type |
| 21 | 9.1 | 12.4 | ND | Neg | G | G | A | Wild-type |
| 22 | 18.9 | 22.7 | ND | Neg | G | G | A | Wild-type |
| 23 | 13.3 | 16.9 | ND | Neg | G | G | A | Wild-type |
| 24 | 25.4 | ND | **29.4** | **Pos** | G | G | **T** | N501Y |
| 25 | 12.8 | 15.9 | ND | Neg | G | G | A | Wild-type |
| 26 | 17.1 | 20.8 | ND | Neg | G | G | A | Wild-type |
| 27 | 17.2 | ND | **21.1** | **Pos** | G | G | **T** | N501Y |
| 28 | 17.7 | ND | **22.1** | **Pos** | G | G | **T** | N501Y |
| 29 | 27.9 | ND | **32.2** | **Pos** | G | G | **T** | N501Y |
| 30 | 22.4 | ND | **26.6** | **Pos** | G | G | **T** | N501Y |
| 31 | 12.7 | ND | **17** | **Pos** | G | G | **T** | N501Y |
| 32 | 13.2 | ND | **18.5** | **Pos** | G | G | **T** | N501Y |
| 33 | 34.90 | 35.8 | ND | Neg | G | G | A | Wild-type |
| 34 | 27.10 | 29.9 | ND | Neg | G | G | A | Wild-type |
| 35 | 19.97 | 22.0 | ND | Neg | G | G | A | Wild-type |
| 36 | 23.65 | 25.9 | ND | Neg | G | G | A | Wild-type |
| 37 | 28.23 | 30.5 | ND | Neg | G | G | A | Wild-type |
| 38 | 24.89 | 28.8 | ND | Neg | G | G | A | Wild-type |
| 39 | 23.13 | 25.6 | ND | Neg | G | G | A | Wild-type |
| 40 | 19.35 | 22.4 | ND | Neg | G | G | A | Wild-type |
| 41 | 24.49 | 26.9 | ND | Neg | G | G | A | Wild-type |
| 42 | 16.30 | 19.1 | ND | Neg | G | G | A | Wild-type |
| 43 | 30.41 | 33.3 | ND | Neg | G | G | A | Wild-type |
| 44 | 21.20 | 23.8 | ND | Neg | G | G | A | Wild-type |
| 45 | 26.96 | 29.7 | ND | Neg | G | G | A | Wild-type |
| 46 | 27.99 | 30.4 | ND | Neg | G | G | A | Wild-type |
| 47 | 25.96 | 28.6 | ND | Neg | G | G | A | Wild-type |
| 48 | 23.29 | 25.8 | ND | Neg | G | G | A | Wild-type |
| 49 | 31.79 | 33.7 | ND | Neg | G | G | A | Wild-type |
| 50 | 20.76 | 23.4 | ND | Neg | G | G | A | Wild-type |
| 51 | 21.56 | 24.8 | ND | Neg | G | G | A | Wild-type |
| 52 | 22.73 | 24.7 | ND | Neg | G | G | A | Wild-type |
| 53 | 17.08 | 19.5 | ND | Neg | G | G | A | Wild-type |
| 54 | 16.34 | 18.8 | ND | Neg | G | G | A | Wild-type |
| 55 | 27.65 | 30.3 | ND | Neg | G | G | A | Wild-type |
| 56 | 18.93 | 21.3 | ND | Neg | G | G | A | Wild-type |
| 57 | 21.52 | 24.3 | ND | Neg | G | G | A | Wild-type |
| 58 | 18.10 | 21.0 | ND | Neg | G | G | A | Wild-type |
| 59 | 16.46 | 19.0 | ND | Neg | G | G | A | Wild-type |
| 60 | 19.09 | 21.7 | ND | Neg | G | G | A | Wild-type |
| 61 | 20.80 | 23.6 | ND | Neg | G | G | A | Wild-type |
| 62 | 21.42 | ND | **23.7** | **Pos** | G | G | **T** | N501Y |
| 63 | 21.7 | 24.7 | ND | Neg | G | G | A | Wild-type |
| 64 | 20.1 | 22.7 | ND | Neg | G | G | A | Wild-type |
| 65 | 17.6 | 20.1 | ND | Neg | G | G | A | Wild-type |
| 66 | 22.8 | 25.7 | ND | Neg | G | G | A | Wild-type |
| 67 | 19.8 | 22.6 | ND | Neg | G | G | A | Wild-type |
| 68 | 19.7 | 22.1 | ND | Neg | G | G | A | Wild-type |
| 69 | 16.8 | 19.3 | ND | Neg | G | G | A | Wild-type |
| 70 | 18.8 | 21.3 | ND | Neg | G | G | A | Wild-type |
| 71 | 19.9 | 22.4 | ND | Neg | G | G | A | Wild-type |
| 72 | 23.6 | ND | **25.6** | **Pos** | G | G | **T** | N501Y |
| 73 | 24.7 | ND | **27.0** | **Pos** | G | G | **T** | N501Y |
| 74 | 22.1 | ND | **24.1** | **Pos** | G | G | **T** | N501Y |
| 75 | 17.0 | ND | **18.8** | **Pos** | G | G | **T** | N501Y |
| 76 | 21.9 | ND | **23.2** | **Pos** | G | G | **T** | N501Y |
| 77 | 24.3 | 26.5 | ND | Neg | G | G | A | Wild-type |
| 78 | 17.33 | ND | **20.12** | **Pos** | G | G | **T** | N501Y |
| 79 | 18.23 | ND | **21.74** | **Pos** | G | G | **T** | N501Y |
| 80 | 22.37 | ND | **24.91** | **Pos** | G | G | **T** | N501Y |
| 81 | 13.54 | 18.98 | ND | Neg | G | A | A | Wild-type |
| 82 | 19.63 | ND | **22.08** | **Pos** | G | G | **T** | N501Y |
| 83 | 23.34 | ND | **25.60** | **Pos** | G | G | **T** | N501Y |
| 84 | 19.65 | ND | **21.96** | **Pos** | G | G | **T** | N501Y |
| 85 | 14.29 | ND | **17.09** | **Pos** | G | G | **T** | N501Y |
| 86 | 16.77 | ND | **19.95** | **Pos** | G | G | **T** | N501Y |
| 87 | 21.30 | ND | **23.09** | **Pos** | G | G | **T** | N501Y |
| 88 | 22.16 | ND | **23.75** | **Pos** | G | G | **T** | N501Y |
| 89 | 20.03 | ND | **22.45** | **Pos** | G | G | **T** | N501Y |
| 90 | 16.44 | ND | **18.46** | **Pos** | G | G | **T** | N501Y |
| 91 | 14.93 | ND | **16.85** | **Pos** | G | G | **T** | N501Y |
| 92 | 15.04 | ND | **16.86** | **Pos** | G | G | **T** | N501Y |
| 93 | 16.38 | ND | **21.15** | **Pos** | G | G | **T** | N501Y |
| 94 | 13.71 | ND | **15.44** | **Pos** | G | G | **T** | N501Y |
| 95 | 17.89 | ND | **19.75** | **Pos** | G | G | **T** | N501Y |
| 96 | 27.11 | 31.80 | ND | Neg | G | G | A | Wild-type |
| 97 | 28.46 | 32.66 | ND | Neg | G | G | A | Wild-type |
| 98 | 20.26 | 24.98 | ND | Neg | G | G | A | Wild-type |
| 99 | 26.55 | 31.17 | ND | Neg | G | G | A | Wild-type |
| 100 | 25.55 | 30.62 | ND | Neg | G | G | A | Wild-type |
| 101 | 20.63 | 25.58 | ND | Neg | G | G | A | Wild-type |
| 102 | 16.87 | ND | **19.57** | **Pos** | G | G | **T** | N501Y |
| 103 | 16.04 | ND | **17.5** | **Pos** | G | G | **T** | N501Y |
| 104 | 13.64 | 15.7 | ND | Neg | G | G | A | Wild-type |
| 105 | 21.50 | 22.9 | ND | Neg | G | G | A | Wild-type |
| 106 | 18.10 | 19.4 | ND | Neg | G | G | A | Wild-type |
| 107 | 18.16 | 19.4 | ND | Neg | G | G | A | Wild-type |
| 108 | 16.52 | 18.6 | ND | Neg | G | G | A | Wild-type |
| 109 | 21.79 | 23.7 | ND | Neg | G | G | A | Wild-type |
| 110 | 20.81 | 22.4 | ND | Neg | G | G | A | Wild-type |
| 111 | 19.32 | 20.9 | ND | Neg | G | G | A | Wild-type |
| 112 | 12.65 | 15.2 | ND | Neg | G | G | A | Wild-type |
| 113 | 21.08 | 23.4 | ND | Neg | G | G | A | Wild-type |
| 114 | 22.37 | 24.2 | ND | Neg | G | G | A | Wild-type |
| 115 | 19.81 | 21.9 | ND | Neg | G | G | A | Wild-type |
| 116 | 18.91 | 20.9 | ND | Neg | G | G | A | Wild-type |
| 117 | 20.64 | 23.6 | ND | Neg | G | G | A | Wild-type |
| 118 | 20.99 | 23.3 | ND | Neg | G | G | A | Wild-type |
| 119 | 17.45 | 19.9 | ND | Neg | G | G | A | Wild-type |
| 120 | 17.30 | ND | **20.0** | **Pos** | G | G | **T** | N501Y |
| 121 | 20.92 | ND | **23.3** | **Pos** | G | G | **T** | N501Y |
| 122 | 15.60 | 17.9 | ND | Neg | G | G | A | Wild-type |
| 123 | 32.45 | 34.2 | ND | Neg | G | G | A | Wild-type |
| 124 | 23.01 | 25.5 | ND | Neg | G | G | A | Wild-type |
| 125 | 25.93 | 28.1 | ND | Neg | G | G | A | Wild-type |
| 126 | 12.08 | 14.5 | ND | Neg | G | G | A | Wild-type |
| 127 | 13.66 | 16.3 | ND | Neg | G | G | A | Wild-type |
| 128 | 12.11 | 14.4 | ND | Neg | G | G | A | Wild-type |
| 129 | 23.63 | 25.4 | ND | Neg | G | G | A | Wild-type |
| 130 | 27.52 | 29.4 | ND | Neg | G | G | A | Wild-type |
| 131 | 24.40 | ND | **26.7** | **Pos** | G | G | **T** | N501Y |
| 132 | 28.29 | **28.70** | **29.3** | **Mix** | MIX<br>base |  |  | MIX base |
| 133 | 22.44 | 23.9 | ND | Neg | G | G | A | Wild-type |
| 134 | 24.06 | 25.6 | ND | Neg | G | G | A | Wild-type |
| 135 | 18.42 | 20.7 | ND | Neg | G | G | A | Wild-type |
| 136 | 17.93 | 19.7 | ND | Neg | G | G | A | Wild-type |
| 137 | 23.90 | ND | **25.4** | **Pos** | G | G | **T** | N501Y |
| 138 | 20.66 | ND | **22.9** | **Pos** | G | G | **T** | N501Y |
| 139 | 27.82 | ND | **30.4** | **Pos** | G | G | **T** | N501Y |
| 140 | 18.76 | ND | **20.8** | **Pos** | G | G | **T** | N501Y |
| 141 | 15.49 | ND | **17.4** | **Pos** | G | G | **T** | N501Y |
| 142 | 15.30 | ND | **17.5** | **Pos** | G | G | **T** | N501Y |
| 143 | 16.07 | ND | **18.5** | **Pos** | G | G | **T** | N501Y |
| 144 | 18.51 | ND | **21.0** | **Pos** | G | G | **T** | N501Y |
| 145 | 14.70 | ND | **17.0** | **Pos** | G | G | **T** | N501Y |
| 146 | 14.11 | ND | **16.7** | **Pos** | G | G | **T** | N501Y |
| 147 | 18.9 | ND | **21.7** | **Pos** | G | G | **T** | N501Y |
| 148 | 20.1 | ND | **22.4** | **Pos** | G | G | **T** | N501Y |
| 149 | 18.7 | ND | **20.6** | **Pos** | G | G | **T** | N501Y |
| 150 | 14.7 | ND | **15.8** | **Pos** | G | G | **T** | N501Y |
| 151 | 11.9 | 14.5 | ND | Neg | G | G | A | Wild-type |
| 152 | 15.0 | 18.3 | ND | Neg | G | G | A | Wild-type |
| 153 | 32.2 | 36.2 | ND | Neg | G | G | A | Wild-type |
| 154 | 23.7 | 27.9 | ND | Neg | G | G | A | Wild-type |
| 155 | 20.8 | 24.1 | ND | Neg | G | G | A | Wild-type |
| 156 | 17.0 | 19.8 | ND | Neg | G | G | A | Wild-type |
| 157 | 34.5 | 36.0 | ND | Neg | G | G | A | Wild-type |
| 158 | 15.1 | 17.8 | ND | Neg | G | G | A | Wild-type |
| 159 | 10.9 | 13.5 | ND | Neg | G | G | A | Wild-type |
| 160 | ND | 37.9 | ND | Neg | G | G | A | Wild-type |

a.ND= Not Detected. Mix means probes for N501 and N501Y were detected when the specimen was run in our assay and the Sanger sequencing results showed signals for both N501 and N501Y.

**Supplementary Table 3:**
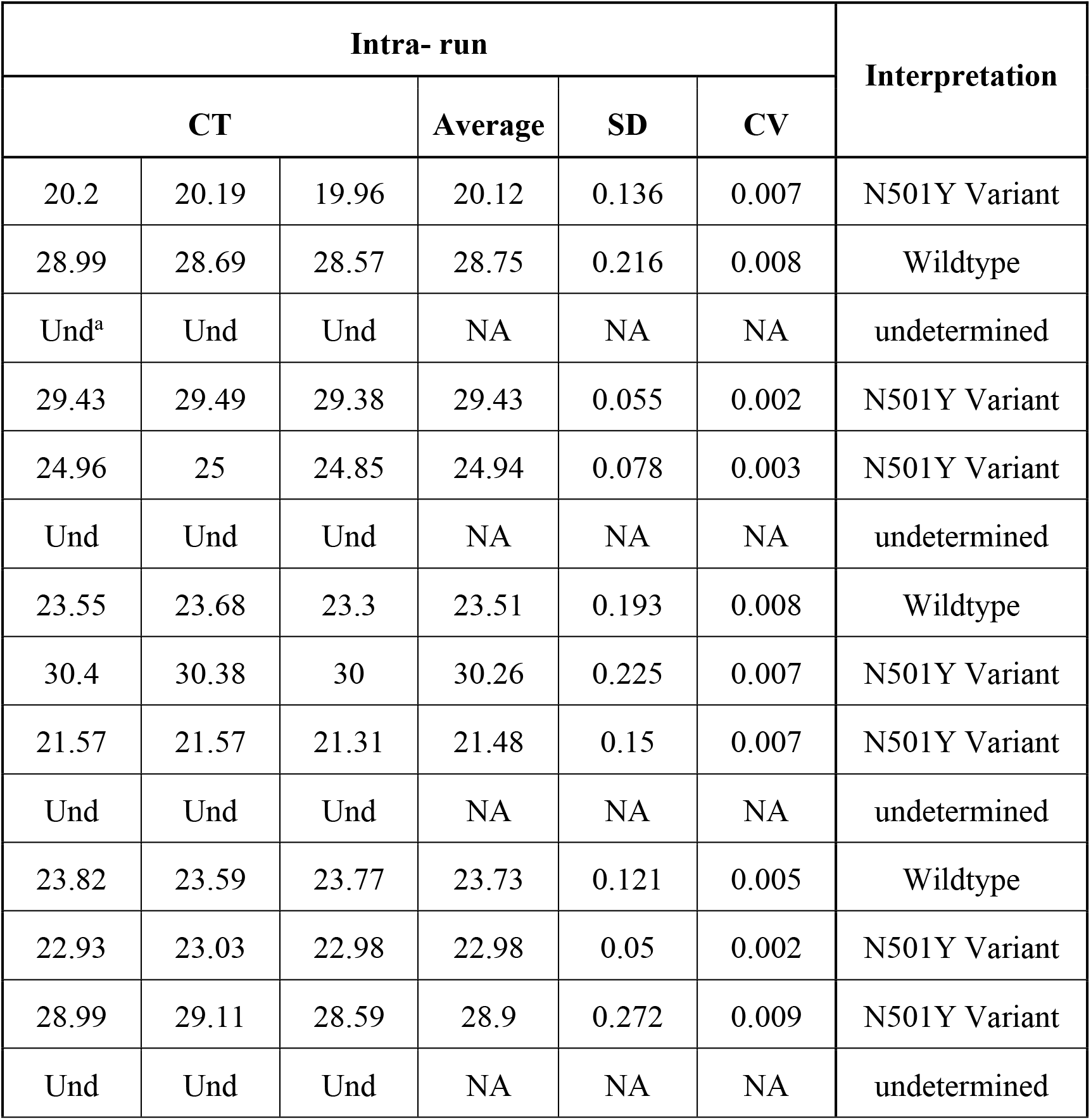
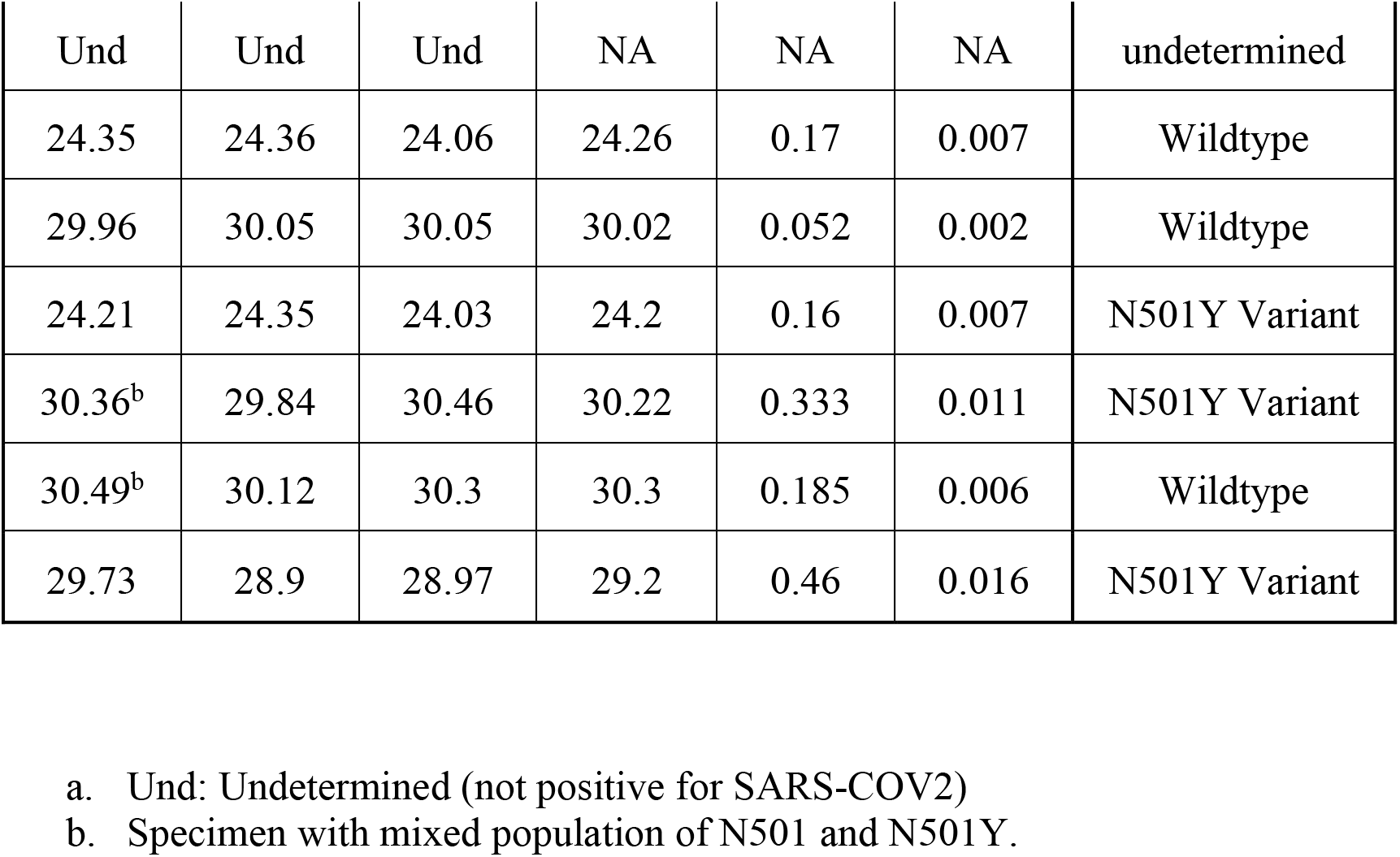
Summary of intra-run repeatability

## Supplementary Appendix A

### PCR Amplification and Sequencing of Partial Spike Gene Methodology

Primers were designed to amplify a 698 bp S gene fragment spanning the region of interest (genome positions: 22516 to 23214). Reverse transcription and amplification of the partial S gene was performed using the OneStep RT-PCR kit (QIAGEN, Hilden, Germany). A 25 µl reaction mixture contained 5 µl of 5X QIAGENOneStep RT-PCR Mix, 1 μl (200 μM) of dNTPs, 1 μl of OneStep RT-PCR enzyme mix, 1 μl of each forward (VOCF) and reverse (VOCR) primers at a final concentration of 0.4 μM (**Supplementary Table 1**), 11μl of water and 5μl of RNA. Reactions were carried out on the Applied Biosystems™ SimpliAmp® Thermal Cycler (Thermo Fisher Scientific, Massachusetts, United States) with the following conditions: 1 cycle at 50 °C for 30 minutes and 1 cycle at 95 °C for 15 minutes, followed by 40 cycles of amplification at 94 °C for 30 seconds, 56 °C for 30 seconds, and 72 °C for 1 minute, and a final extension cycle of 72 °C for 10 minutes.

Applied Biosystems™ BigDye™ Terminator v3.1 Cycle Sequencing Kit (Thermo Fisher Scientific, Massachusetts, United States) was used for bidirectional sequencing of PCR fragments following manufacturer’s recommendations. Each reaction contained 0.5 µl of Terminator Ready Reaction Mix, 3.75 µl of 5X buffer, 1µl of primer mix and 1 µl amplified PCR product, and 13.75μl of water in a final volume of 20 µl. Default cycling conditions for the BigDye™ Terminator reaction were used as follows: 96°C for 1 minute followed by 35 cycles of 96°C for 10 seconds, 50°C for 5 seconds, and 60°C for 4 minutes. BigDye™ reaction products were purified using an EDTA/ ethanol precipitation method and resuspended in 20µl Hi-Di™ Formamide (Thermo Fisher Scientific, Massachusetts, United States). Nucleotide sequences were determined using the Applied Biosystems™ ABI 3130xl Genetic Analyzer (Thermo Fisher Scientific, Massachusetts, United States) and analyzed using FinchTV software (Geospiza, Inc.).

## References

1. COVID-19 Weekly epidemiological update. World Health Organization. Published February 25, 2021. https://www.who.int/publications/m/item/covid-19-weekly-epidemiological-update

2. Rambaut A, Loman N, Pybus O, et al. Preliminary genomic characterisation of an emergent SARS-CoV-2 lineage in the UK defined by a novel set of spike mutations. Virological. Published December 2020. https://virological.org/t/preliminary-genomic-characterisation-of-an-emergent-sars-cov-2-lineage-in-the-uk-defined-by-a-novel-set-of-spike-mutations/563.

3. Tegally H, Wilkinson E, Giovanetti M, et al. Emergence and rapid spread of a new severe acute respiratory syndrome-related coronavirus 2 (SARS-CoV-2) lineage with multiple spike mutations in South Africa. medRxiv. 2020. http://doi.org/10.1101/2020.12.21.20248640.

4. Faria NR, Claro IM, Candido D, et al. Genomic characterisation of an emergent SARS-CoV-2 lineage in Manaus: preliminary findings. Virological. Published Jan 12, 2021. Accessed February 6, 2021. https://virological.org/t/genomic-characterisation-of-an-emergent-sars-cov-2-lineage-in-manaus-preliminary-findings/586.

5. Volz E, Mishra S, Chand M, et al. Transmission of SARS-CoV-2 Lineage B.1.1.7 in England: Insights from linking epidemiological and genetic data. medRxiv. 2021. https://doi.org/10.1101/2020.12.30.20249034.

6. Davies NG, Barnard RC, Jarvis CI, Kucharski AJ, Munday J, Pearson CAB, et al. Estimated transmissibility and severity of novel SARS-CoV-2 variant of concern 202012/01 in England. medRxiv. 2020. https://www.medrxiv.org/content/10.1101/2020.12.24.20248822v1

7. Horby P, Huntley C, Davies N, Edmunds J, Ferguson N, Medley G, Semple C. NERVTAG note on B.1.1.7 severity. GOV.UK. Published January 21, 202. https://assets.publishing.service.gov.uk/government/uploads/system/uploads/attachment_data/file/961037/NERVTAG_note_on_B.1.1.7_severity_for_SAGE_771_.pdf

8. B.1.1.7. SARS-CoV-2 Lineages. Published February 20, 2021. https://cov-lineages.org/global_report_B.1.1.7.html

9. Daily epidemiologic summary: COVID-19 in Ontario: January 15, 2020 to April 5, 2021. Ontario Agency for Health Promotion and Protection (Public Health Ontario). https://www.publichealthontario.ca/-/media/documents/ncov/epi/covid-19-daily-epi-summary-report.pdf?la=en

10. Risk Assessment: Risk related to the spread of new SARS-CoV-2 variants of concern in the EU/EEA-first update. European Centre for Disease Prevention and Control. Published January 21, 2021. https://www.ecdc.europa.eu/en/publications-data/covid-19-risk-assessment-spread-new-variants-concern-eueea-first-update

11. Wibmer CK, Ayres F, Hermanus T, Madzivhandila M, Kgagudi P, Oosthuysen B, et al. SARS-CoV-2 501Y.V2 escapes neutralization by South African COVID-19 donor plasma. Nat Med. 2021;https://doi.org/10.1038/s41591-021-01285-x

12. Comprehensive mapping of mutations to the SARS-CoV-2 receptor-binding domain that affect recognition by polyclonal human serum antibodies. Cell Host & Microbe. 2021; 29(3):463–476.

13. B.1.351. SARS-CoV-2 Lineages. Published February 20, 2020. https://cov-lineages.org/global_report_B.1.351.html

14. Sabino EC, Buss LF, Carvalho MPS, Prete Jr CA, Crispim MAE, Fraiji NA, et al. Resurgence of COVID-19 in Manaus, Brazil, despite high seroprevalence. The Lancet. 2021; 6736(21)00183-00185. https://doi.org/10.1016/ S0140-6736(21)00183-5.

15. P.1. SARS-CoV-2 Lineages. Published February 20, 2021. https://cov-lineages.org/global_report_P.1.html

16. SARS-CoV-2 (COVID-19 Virus) Variant of Concern (VoC) Surveillance. Ontario Agency for Health Protection and Promotion (Public Health Ontario). https://www.publichealthontario.ca/en/laboratory-services/test-information-index/covid-19-voc

17. Bal A, Destras G, Gaymard A, Stefic K, Marlet J, Eymieux S, et al. Two-step strategy for the identification of SARS-CoV-2 variant of concern 202012/01 and other variants with spike deletion H69–V70, France, August to December 2020. Eurosurveillance. 2021;26(3):pii=2100008. https://doi.org/10.2807/1560-7917.ES.2021.26.3.2100008

18. Vogels CBF, Breban M, Alpert T, Petrone ME, Watkins AE, Hodcroft EB, et al. PCR assay to enhance global surveillance for SARS-CoV-2 variants of concern. medRxiv, 2021. https://doi.org/10.1101/2021.01.28.21250486.

19. Durner J, Burggraf S, Czibere L, Tehrani A, Watts DC, Becker M. Fast and cost-effective screening for SARS-CoV-2 variants in a routine diagnostic setting. Dental Materials. 2021. https://doi.org/10.1016/j.dental.2021.01.015.

20. Summary report: SARS-CoV-2 variants of concern: results of point-prevalence study. Ontario Agency for Health Protection and Promotion (Public Health Ontario). https://www.publichealthontario.ca/-/media/documents/ncov/voc/2021/02/sars-cov-2-variants-point-prevalence.pdf?la=en

21. B.1.525. SARS-CoV-2 Lineages. Published February 20, 2021. https://cov-lineages.org/global_report_B.1.525.html

22. Corman VM, Landt O, Kaiser M, Molenkamp R, Meijer A, Chu DKW, et al. Detection of 2019 novel coronavirus (2019-nCOV) by real-time RT-PCR. Eurosurveillance. 2020;25(3):pii=2000045. https://doi.org/10.2807/1560-7917.ES.2020.25.3.2000045

